# Understanding Telomere Biology in Hematopoietic Cell Transplantation: A Dynamical Systems Perspective

**DOI:** 10.1101/2025.01.20.24319630

**Authors:** Amir A. Toor, Morgan Horton, Haniya Khalid, Elizabeth Krieger, Tsung-Po Lai, Stephen R. Spellman, John E. Levine, Wael Saber, Abraham Aviv, Valerie Stewart, Shahinaz M Gadalla

## Abstract

**Background.:** T cell proliferation and repertoire reconstitution is a hallmark of successful hematopoietic cell transplantation (HCT). This process may be modeled as a dynamical system and in such a system, precise telomere length (TL) measurement may reflect the proliferative capacity of donor T cells. TL for different chromosomes span a few orders of magnitude, and different T cell clones will display variable TL; these differences across the population are not represented when examining average TL. This study aims to develop a method that integrates the entire spectrum of TL observed within a sample to better understand the influence on clinical outcomes following HCT.

**Methods.:** To better reflect the entire span of TL , we used data generated using the TeLSA PCR technique that provided discrete measurments of individual telomeres for each DNA sample for 72 stem cell transplant (SCT) donor-recipient pairs. Donor and recipient TeSLA TL measurements was performed on samples taken before and 90 days post HCT, respectively. Set correspondence mathematical techniques and area under the curve (AUC) calculations were used to measured donor-recipient TL differences (delta-TL) incorporating the full distribution of measured TL from each sample.

**Results.:** Telomere band lengths ranged from 350 basepairs (BP) to 16.7 kilobases with a logarithmically declining distribution in all samples when arrayed in a descending order. Set correspondence methods yielded TL averages which were highly correlated with AUC calculations (r >0.9, p<0.001 for all) The *AUC delta-TL* method predicted patient overall survival (P-log rank <0.0001). HCT recipients with intermediate degrees of telomere attrition (25^th^-75^th^ percentile) post-HCT experienced the best outcomes (2 years overall survival; OS=92%), whilst donors with the least (<25^th^ percentile; 2 years OS=33%; adjusted HR *vs.* intermediate shortening=9.3, p=0.001) and the greatest (>75^th^ percentile; 2 years OS=59%; adjusted HR=6.0, p=0.01) shortening had worse outcomes. By contrast, using the traditional method based on donor-recipient difference in TeSLA mean telomere length did not demonstrate survival association in this small sample set (p log-rank=0.95).

**Conclusion.:** The findings described herein suggest that the degree of donor telomere attrition may reflect T cell proliferation and alloreactivity following transplant. Accounting for the entire span of telomere lengths, could better identify post-transplant risk groups.

## Introduction

Contemporary allogeneic hematopoietic cell transplantation (HCT) has evolved towards utilization of younger stem cell donors^1^, sometimes at the expense of HLA matching^2^. This shift is driven by the improved clinical outcomes observed following HCT from younger donors^1^. Studies show improved T cell repertoire reconstitution^3^, less graft versus host disease (GVHD)^4^ and superior relapse protection^5^ when younger donors are utilized. This could be because of a more polyclonal and less ‘exhausted’ T cell repertoire, a more favorable ratio of naïve to memory T cells in younger individuals^6^ and possibly a greater proliferative capacity of both hematopoietic progenitors and T cells ^7^. While the first two have been studied extensively, the last of these remains more speculative. Nevertheless, proliferative capacity may be indirectly evaluated objectively by measuring telomere lengths (TL). Telomeres are thymidine-adenine-guanine hexamer repeat sequences (TTAAGG)^1^ found at the 3’- (telomeric) ends of chromosomes. The length of these repeats is generally proportional to the length of the chromosomes they protect^8^. With age, TL in somatic cells, including hematopoietic cells, shorten by 30–35 bp per year. TL attrition is accelerated by events such as HCT^9–11^. Recent studies showed associations between longer donor TL and better HCT survival in patients with early-stage leukemia/MDS or severe aplastic anemia, respectively.

While TL measurement should be a logical choice for assessing donor suitability for transplantation, the clinical application of this method has not been straightforward. Studies trying to assess the impact of donor TL and variation in TL following SCT have been challenged by inconsistencies because of assay limitations, particularly for the most commonly used qPCR method. Among precise assays for TL measurements are Southern blotting, flow FISH, and more recently **<u>Te</u>**lomere **<u>S</u>**hortest **<u>L</u>**ength **<u>A</u>**ssay (TeSLA)^12^, all of which express TL in mathematical averages or median values that are then evaluated for their association with clinical outcomes. Nevertheless, biomarker impact on clinical outcomes is context dependent. Telomere length alteration in T cells will be manifest when there are alloreactive T cell clones, defined by T cell receptor sequences, proliferating in response to allo-antigen in the context of GVHD or responding to other antigenic pressures (**Appendix I**); it will be the telomeres in these proliferating T cell clones that will shorten, with TL being preserved in other non-proliferating clones. In the recipient of HCT from a young donor, large scale TL attrition may not be readily observed due to averaging over many T cell clones, especially when TL itself is being averaged for analysis. T cell clonal growth has been modelled as a dynamical system where the T cell repertoire constitutes a multi-component vector which transforms with respect to T cell clonal dominance following transplantation^13–15^. Logically, this clonal repertoire (vector) transformation ought to be reflected in TL attrition observed after transplantation and be applicable to understanding HCT outcomes. However, this requires more precise TL measurement than some of the current techniques are able to provide.

Compared to all other TL measurement methods, TeSLA provides a full distribution of TL per DNA content (ranging between few hundred bp to 20 kb) and is powered by its ability to detect and quantify the shortest telomeres, which are believed to be the trigger for cellular senescence and apoptosis^16,17^. In all previous HCT TL studies, including those using TeSLA^18^, difference in donor-recipient mean TL (delta-TL) was used to describe post-HCT changes and correlate them with clinical outcomes. However, these studies were limited since the delta-TL reported were averaged over two different distributions, potentially missing full picture of post-HCT TL changes. The current study was undertaken to develop a method that integrates the entire spectrum of TL observed in any given sample and to better understand its influence on clinical outcomes following HCT.

## Methods

De-identified, detailed numeric blood leukocyte TeSLA TL measurement data from 72 allogeneic HCT donor-recipient pairs (DRP) from a previously-published study^18^ were analyzed (**Supplementary Table 1**). Donor samples were collected before stem cell mobilization or collection as part of the CIBMTR Biorepository Protocol^19^(https://cibmtr.org/CIBMTR/Resources/Biorepository-Inventories). Recipient samples were collected at three months post-transplant (mean=90 +/-14 days) from patients enrolled on the BMT CTN 1202 Protocol^20^(https://bmtctn.net/study-proposals). The data analyzed included band lengths of telomeres measured by TeSLA (mean number of bands=129 per sample) that were determined as previously described.^2^

The magnitude of TL shortening post-HCT (delta-TL) in original study was calculated by subtracting TeSLA software output of the arithmetic mean of TL in recipient post-HCT from that of their donor . This is referred to herein as the *original delta-TL* method. In the present analysis, delta-TL calculation took into account individual telomere bands to incorporate the entire spectrum of TL observed. To accomplish this, a set correspondence method was utilized in which the TL (band length in bp) were ordered in *single column matrices* in descending order for donors and recipients. These column matrices represented the range of TL measured in each sample, comprising unique *sets* describing the state of each system at the time of measurement. Through examination of these data *sets* in their entirity, the TL of both the proliferating and non-proliferating T cell clones is reflected in the analysis while the mean TL approach is influenced by the more numerous shorter TL in these populations. The set correspondence method measured the delta-TL of the closest matched bands.

To determine the change from baseline donor telomere state to the recipient telomere state at three months, the difference between the two column matrices was calculated, measuring the *transformation* of these matrices (sets of TL) that occurred through the course of transplantation. This was accomplished by considering both matrices as sets of TL values and subtracting the corresponding elements of the sets. This set correspondence operation was performed using two methods. In the first analaysis, a 1:1 *correspondence* was established where each (*i^th^*) element of the donor (*Di*) set was mapped to a corresponding (*j^th^*) element in the recipient (*Rj*) set. If there were more elements in either of the sets than the other, a value of zero was used in the set with fewer elements to correspond to the extra elements in the larger set (see **Supplementary Method Appendix II**). In a second analysis, a *surjection function* was utilized to avoid the introduction of zero in the matrix with fewer measurements (bands). For this calculation, the assumptions that donor TL will always be longer than recipient TL and that TL of 0 and a delta comprised of a negative number is most likely inconsistent with normal physiology were made. Using this approach, in instances of unequal set size, longer donor sequences could be mapped to one or more equal or shorter recipient sequences until all recipient sequences had been mapped (**Supplementary Figure 1**). The distribution of the delta-TL values for each DRP was then summarized by geometric means for the calculated delta with surjection (*geo delta-TL*) and median for the 1:1 correspondence deltas (*median delta-TL*).

To avoid loss of information through reliance on central values, an area under the curve calculation (*AUC delta-TL*) was performed by plotting the individual band lengths in each sample (donor and recipient) as a function of the number of bands in that sample. AUC was determined using the trapezoidal rule to give an AUC measure for each sample. This integration rule divides the section under the telomere band length curve into multiple trapezoids of equal width on the x-axis and uses the area of a trapezoid fomula [*A= ½(a+b)(h)*] to calculate their individual areas. The individual areas were then summed to estimate the total area under the curve (**Supplementary Figure 2**).

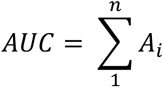

The AUC calculations were confirmed using the programming platform MATLAB® (*MathWorks, Natick, MA*). ^3^ The difference between the AUC of the donors and recipients in each DRP, *AUC delta-TL_D-R_* were then calculated as follows:

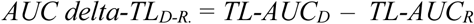

## Statistical analysis

Pearson correlation coefficient was used to test the correlations between delta-TL calculated by both the original and new methods. Patient characteristics and transplant factors were compared across different categories of delta-TL using chi-square or Fisher exact test for categorical variables and t-test for continuous variables. Delta-TL categories were categorized as least shortening (*<25^th^ percentile*), most shortening(*>75^th^ percentile*), and intermediate shortening (*25-75^th^ percentile)*.

The Kaplan-Meier method was used to calculate the probabilities of survival across the three delta-TL groups noted above and statistical differences were tested using the log-rank test. Follow-up started 90 days after HCT. Cox proportional hazard model was used to calculate hazard ratios (HR) and 95% confidence intervals (CI) of all-cause mortality for the least and most delta-TL shortening compared with the intermediate group. Models were adjusted for potential confounders (statistically significant variables in **Table 1**).

**Table 1.** The correlation coefficient between different measurements of telomere length difference between donors and recipients

|  | Delta-TL | Geo Delta-TL | Med Delta-TL | AUC Delta-TL |
| --- | --- | --- | --- | --- |
| Delta-TL | 1 | 0.53 | 0.58 | 0.56 |
| Geo Delta-TL | 0.53 | 1 | 0.95 | 0.94 |
| Med Delta-TL | 0.58 | 0.95 | 1 | 0.98 |
| AUC Delta-TL | 0.56 | 0.94 | 0.98 | 1 |

\*p-value <0.001 for all

## Results

### Telomere band length distribution

Telomere band lengths when plotted in a descending order followed a non-linear, logarithmic distribution (**Figure 1**), i.e., longer bands tended to be spaced farther apart, as opposed to the shorter ones, where the difference between the successive bands was smaller and these were more tightly spaced. These patterns were consistently observed across all the donor-recipient samples examined and is consistent with the notion that TL distribution may be mathematically described since it varies as a function of the ubiquitous growth constant, *e*. **Supplemental Figure 3**, b and distribution slopes were different between individuals (varied exponents).

**Figure 1.**
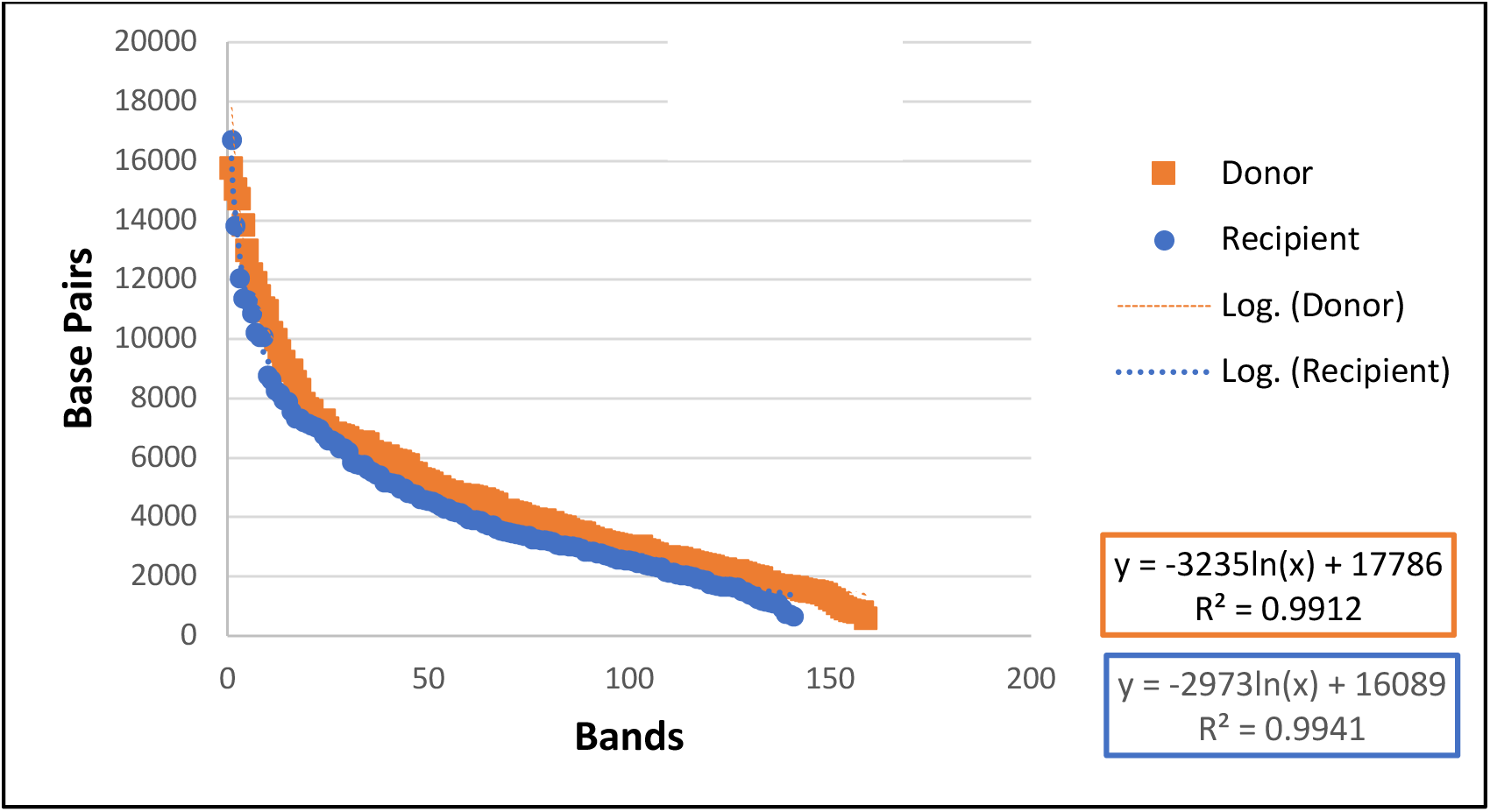
Telomere band lengths plotted in descending order for demonstrated logarithmic decline with variable magnitudes of telomere attrition recipient (in orange) and matched donor (in blue). TL in base-pairs on the y-axis, successive bands on the x-axis.

### Donor-recipient telomere length change (delta-TL) post-HCT

Using the set correspondence rules, TL demonstrated marked variability across all DRP, with different slopes of decline in the telomere length attrition curves and significant variability in the consequent measures of difference, such as *AUC delta-TL* (p paired t-test <0.0001; **Figure 2**). This suggested a varied evolution of telomeric state that the blood cells acquired in different DRP. All new delta-TL methods (set correspondence rules and AUC) demonstrated excellent correlation between one another (correlation coefficient r >0.9, p <0.0001), but the correlations of these methods with the original delta-TL, calculated by subtracting the arithmetic mean of recipient TL from the mean of the donor were weaker (r <0.6, p <0.001; **Table 1**). **Supplemental Table 2** summerizes the distribution of delta-TL by different calculation methods.

**Figure 2.**
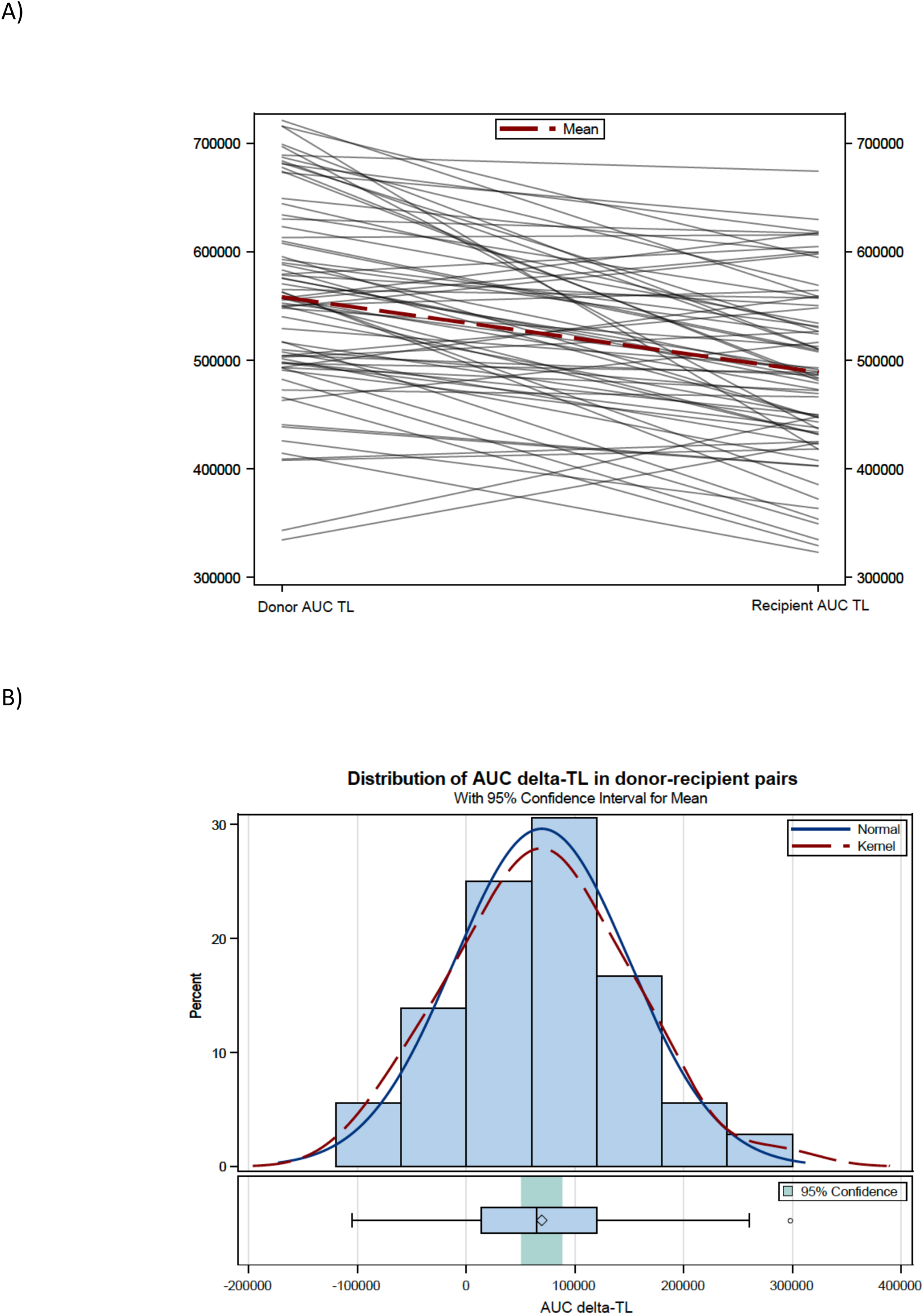
Donor and recipient AUC-TL. A) spegetti plot with lines connecting corresponding values for donor and recipient AUC TL in each DRP. B) Distribution of AUC detla-TL across the entire cohort

### Factors associated with delta-TL variation

Older donor age was associated with lowest delta-TL shortening in all three methods (**Figure 3** and **Supplemental Figure 4**). Delta-TL categories by the three tested methods were associated with donor type, with significantly greater TL shortening in HLA-matched unrelated donors compared to sibling donor-recipient pairs (p<=0.05). Lowest TL shortening by *AUC delta-TL* was associated with severe acute GVHD (aGVHD; p=0.05), while peripheral blood stem cell grafts were associated with greatest TL shortening by *geo delta-TL* and *median delta-TL* (p <0.05). No such associations were observed using the original method of TL attrition calculation in the same DRP set (**Table 2** and **Supplemental Table 3**).

**Figure 3.**
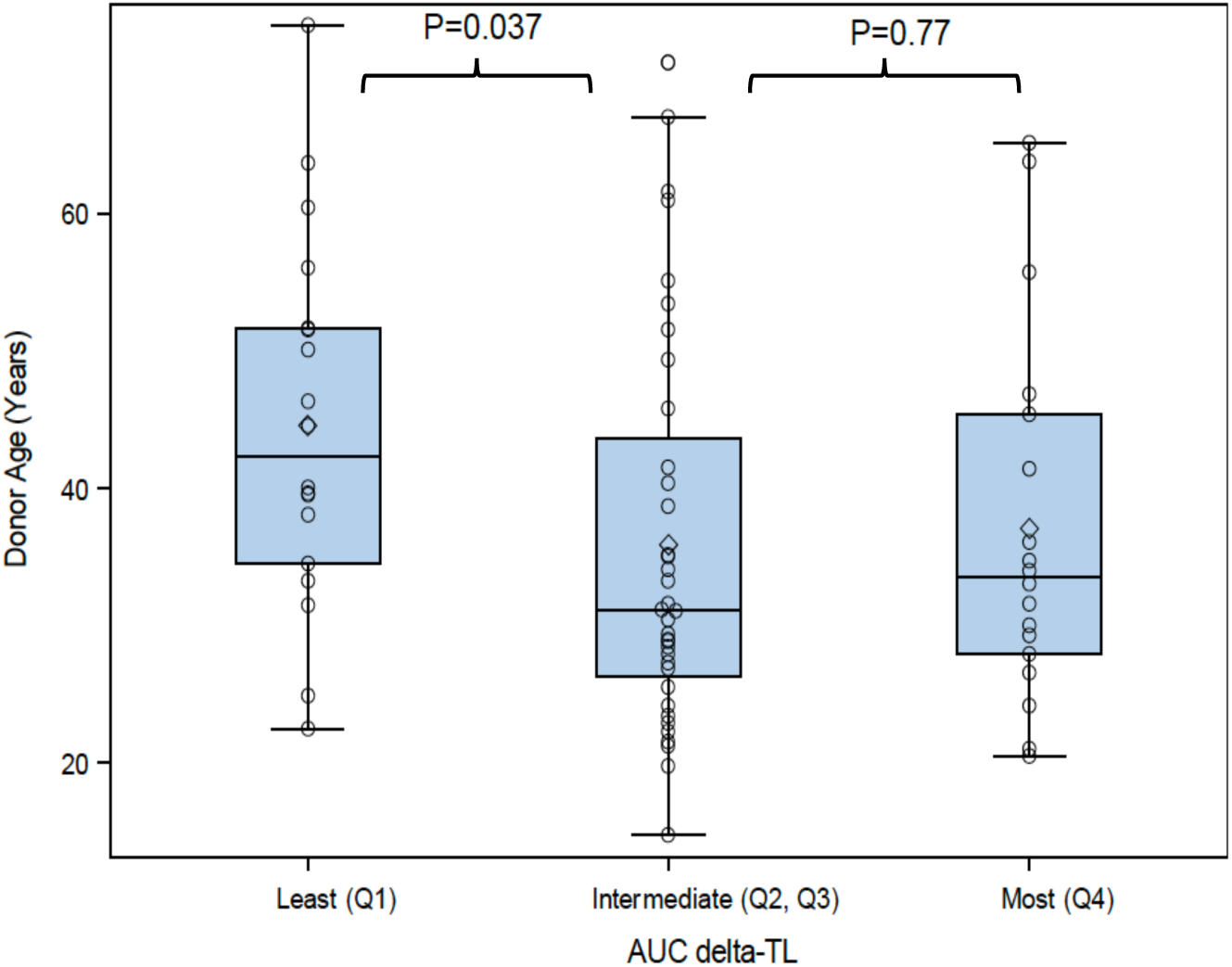
The impact of donor age on *AUC delta-TL*. Younger donors with intermediate TL attrition.

**Table 2.** Factors Associated with *AUC delta-TL*

|  | Least shortening<br>(N=18) | Intermediate (N=36) | Most shortening (N=18) | P<sup>1</sup> |
| --- | --- | --- | --- | --- |
| **Patient age**<br>**(mean, SD)** | 49 (15.5) | 48.4 (19.1) | 52 (15) | 0.71 |
| **Donor age**<br>**(mean, SD)** | 44.6 (13.5) | 35.9 (14.3) | 37.1 (13.5) | 0.09 |
|  | N (%) |  |  |  |
| **Recipient sex** |  |  |  | 1.0 |
| **Male** | 11 (61.1) | 22 (61.1) | 11 (61.1) |  |
| **Female** | 7 (38.9) | 14 (38.9) | 7 (38.9) |  |
| **Donor Sex** |  |  |  | 0.84 |
| **Male** | 10 (55.6) | 17 (47.2) | 9 (50) |  |
| **Female** | 8 (44.4) | 19 (52.8) | 9 (50) |  |
| **Disease** |  |  |  | 0.54 |
| **Myeloid malignancy** | 12 (66.7) | 21 (58.3) | 13 (72.2) |  |
| **Lymphoid malignancy** | 3 (16.7) | 12 (33.3) | 4 (22.2) |  |
| **Other<sup>2</sup>** | 3 (16.7) | 3 (8.3) | 1 (5.6) |  |
| **KPS** |  |  |  | 0.37 |
| **90-100** | 20 (55.5) | 7 (39.0) | 11 (61.1) |  |
| **70-80** | 16 (45.5) | 11 (61) | 7 (30.9) |  |
| **Donor type** |  |  |  | **0.02<sup>3</sup>** |
| **Matched sibling** | 11 (61.1) | 11 (30.5) | 3 (16.7) |  |
| **Unrelated** | 6 (33.3) | 24 (66.7) | 12 (66.6) |  |
| **Haplo-identical** | 1 (5.6) | 1 (2.8) | 3 (16.7) |  |
| **Graft source** |  |  |  | 0.59 |
| **Bone marrow** | 4 (22.2) | 8 (22.2) | 2 (11.1) |  |
| **Peripheral blood** | 14 (77.8) | 28 (77.8) | 16 (88.9) |  |
| **GVHD prophylaxis** |  |  |  | 0.45<sup>3</sup> |
| **Tacro-based** | 11 (61.1) | 28 (77.8) | 15 (83.3) |  |
| **CSA-based** | 2 (11.1) | 3 (8.3) | 0 (0) |  |
| **Other** | 5 (27.8) | 5 (13.9) | 3 (16.7) |  |
| **Regimen intensity** |  |  |  | 0.92 |
| **Myeloablative** | 10 (55.6) | 23 (63.9) | 12 (66.7) |  |
| **RIC** | 6 (33.3) | 9 (25.0) | 5 (27.8) |  |
| **NMA** | 2 (11.1) | 4 (11.1) | 1 (5.5) |  |
| **Recipient-Donor CMV serostatus** |  |  |  | 0.71 |
| **Both negative** | 3 (17.6) | 11 (30.5) | 6 (33.3) |  |
| **Both positive** | 10 (58.8) | 13 (36.1) | 6 (33.3) |  |
| **Negative-Positive** | 2 (11.8) | 4 (11.1) | 3 (16.7) |  |
| **Positive-Negative** | 2 (11.8) | 8 (22.2) | 3 (16.7) |  |
| **Acute GVHD (2-4)** |  |  |  | 0.30 |
| **Yes** | 8 (44.4) | 14 (38.9) | 11 (61.1) |  |
| **No** | 10 (55.6) | 22 (61.1) | 7 (38.9) |  |
| **Acute GVHD (3-4)** |  |  |  | 0.05<sup>3</sup> |
| **Yes** | 5 (27.8) | 3 (8.3) | 6 (33.3) |  |
| **No** | 13 (72.2) | 33 (91.7) | 12 (66.7) |  |

<sup>1</sup>chi-square or t-test unless otherwise specified
<sup>2</sup> Other acute leukemia, not specified, severe aplastic anemia, and immune disorders
<sup>3</sup> Fisher exact test

### Impact of delta-TL attrition on clinical outcomes

Focusing on the *AUC delta-TL* method, overall survival (OS) was highest in the patients who had intermediate shortening in their TL (3 year OS=92%) compared with those with greatest shortening (OS=59%) and those with least shortening, (OS=33%, p log-rank <0.0001;**Figure 4**). Multivariable models adjusted for donor age (continuous), donor type (related or unrelated), and incidence of aGVHD (Yes or No) verified the impact of *AUC delta-TL* on patients on OS. Compared to patients with intermediate shortening in *AUC delta-TL*, the HR of all-cause mortality in patients with least shortening was 11.2, (95%CI 2.6-47.3, p=0.001), and that for greatest shortening was 6.8 (95%CI 1.6-27.7, P=0.007; **Table 3**). Similar observations, but weaker non-statistically significant HR, were noted with *geo delta-TL* and *median delta-TL*, but no OS association with the original method of delta-TL calculation was seen (**Supplementary Table 4**).

**Figure 4.**
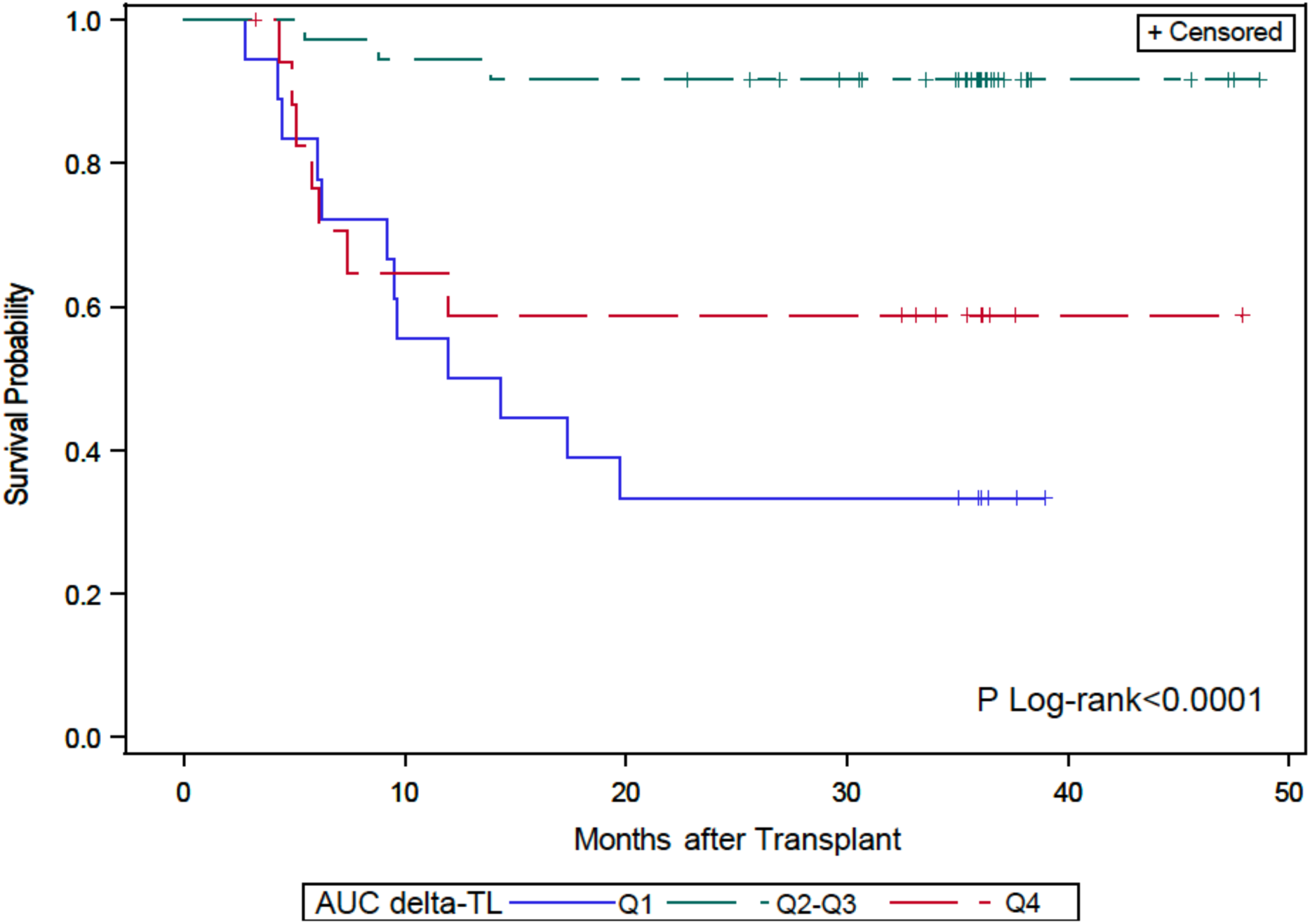
Overall survival in HCT recipients by magnitude of TL attrition calculated as *AUC delta-TL*. Patients with least and greatest TL attrition experience worse outcomes.

**Table 3.**
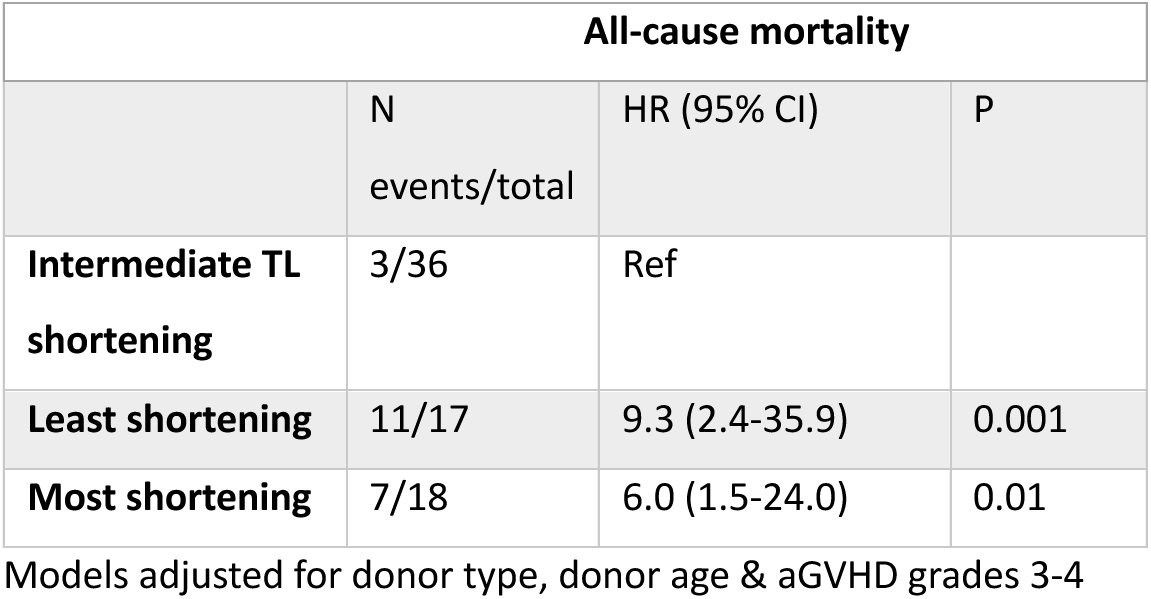
Adjusted association between *AUC delta-TL* and post-HCT mortality of any cause

| All-cause mortality |  |  |  |
| --- | --- | --- | --- |
|  | N<br>events/total | HR (95% CI) | P |
| **Intermediate TL shortening** | 3/36 | Ref |  |
| **Least shortening** | 11/17 | 9.3 (2.4-35.9) | 0.001 |
| **Most shortening** | 7/18 | 6.0 (1.5-24.0) | 0.01 |

Models adjusted for donor type, donor age & aGVHD grades 3-4

Causes of death in DRP in the different delta-TL shortening subgroups were asymmetrically represented, with the majority of relapse death happening in those with least shortening (N=7/12; 58.3%) and most of the GVHD-related death happening in those with greatest shortening (N=3/5; 60%; **Supplementary Table 3**).

## Discussion

Allogeneic HCT is a complex regenerative process where a variety of different hematopoietic progenitors reconstitute at varying rates depending on pharmacotherapy administered to prevent GVHD and promote engraftment. Indeed, hematopoietic cell populations recover at varying rates and with noted variation in TL. Younger donors are more apt to have longer telomeres and, given that these are in many instances unrelated donors selected for older transplant recipients, are more likely to demonstrate antigen-driven T cell proliferation. Young donors might thus demonstrate a greater degree of telomere attrition if alloreactive T cell clones represent a measurable circulating population. It is this notion that the findings of this study support: firstly, that the greatest measure of telomere attrition was seen in recipients of unrelated donor transplants, and secondly the magnitude of TL attrition is germane to allograft outcomes. More broadly, the TL distribution in unfractionated blood samples, as measured by TeSLA is not linear, nor randomly distributed, but rather it declines as a logarithmic function of the number of bands observed. In other words, telomeres elongate during DNA replication as a function of the growth constant *e*.

Transplant outcomes are inherently dependent on donor engraftment and immune recovery. The latter is responsible for protection from opportunistic infections and disease relapse, but conversely might cause or contribute to GVHD. Aligned with these principles is the study’s striking finding that the survival advantage observed in those recipients who had intermediate degree of telomere attrition over the first 90 ± 14 days following transplantation. This degree of telomere shortening was associated with less relapse than limited shortening. Limited TL shortening was associated with older donor age, suggesting a lack of alloreactive T cell proliferation. Additionally, the higher magnitude of TL shortening was also associated with excess mortality risk post-HCT in the present study, possibly through adversely influencing the risk of GVHD as well as the mortality related to GVHD. Thus, alloreactivity driven T cell proliferation woud yield a greater degree of telomere shortening and *vice versa*, which possibility is reflected in the correlation of telomere length with clinical outcomes observed here (**Figure 5**). In the absence of accompanying T cell data to correlate with these TL observations these associations, while statistically significant, remain mechanistically speculative, but biologically plausible.

**Figure 5.**
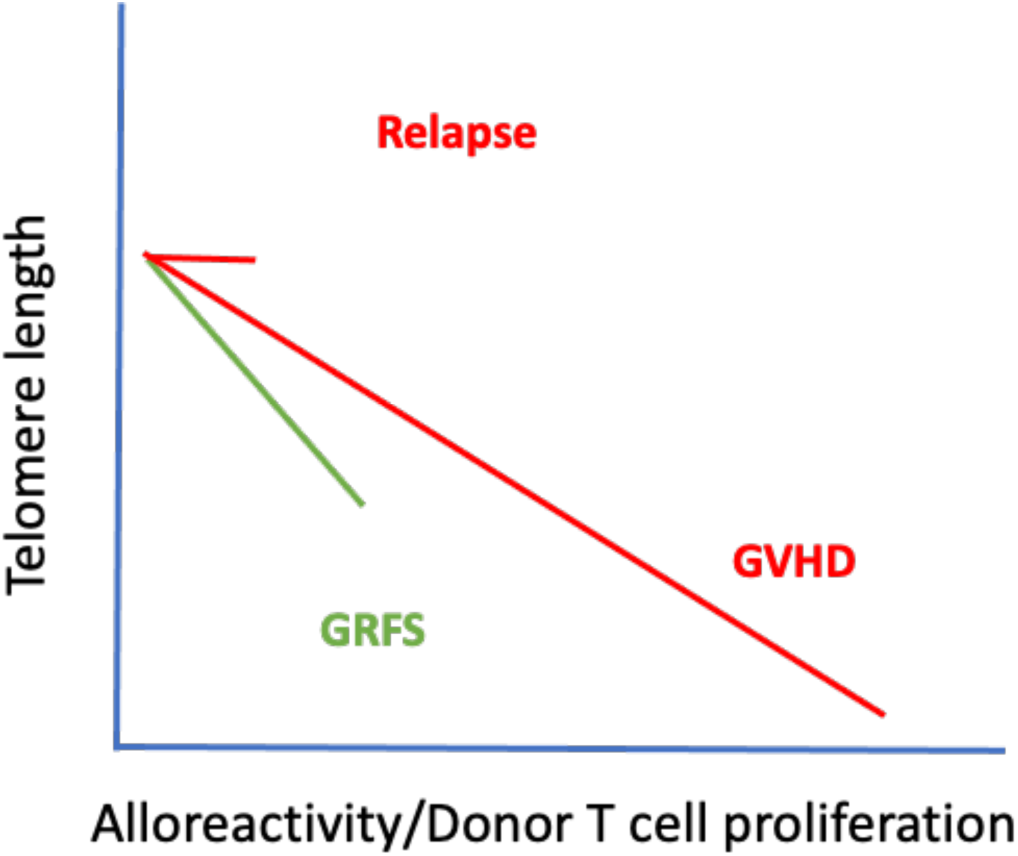
Schematic of the proposed effect of alloreactivity-driven donor T cell proliferation on delta-telomere length, and the subsequent correlation of telomere length with clinical outcomes following allogeneiec HCT. This graphic depicts donors with equal telomere length at outset, and the same span of time.

An important novelty of this study is the method of calculating delta-TL variation in individual DRP. Conventional methods of analyzing TeSLA TL data have involved taking a difference between arithmetic means of values observed for donor and recipient TL, or quantifying the frequency of shortest TL which are deterministic of cell proliferation. This notion would have the fate of the cells hinge on the shortest chromosomes, and seems implausible because the length of telomeres is proportional to the length of chromosomes. Moreover, the observation that telomeres decline according to a logarithmic function relative to their number, spanning at least three orders of magnitude from 10² to 10⁴, suggests that using arithmetic means for measurement could introduce bias. This is because there is a disproportionately larger number of shorter telomere bands compared to longer ones, which skews the mean. Consequently, calculations based on these non-representative averages could be flawed, potentially complicating the identification of clinical associations. In support of this point, while using conventional method to calculate delta-TL in this set did not show associations with clinical outcomes; yet, a similar association between limited TL attrition and risk of relapse or death was detected in a larger sample set of 384 patients (delta-TL was calculated as the difference between arithmetic mean in DRP with TL measured by the precise Southern blot assay)^21^. This indicates that mathematical methods used in the current analysis in which the full distribution of TL in the DRP was considered in the delta-TL calculations led to a higher resolution of estimates that was much more efficient in detecting clinical associations.

The results reported herein, though from a small and clinically varied dataset, are consistent with known transplant immunobiology principles, such as transplants from young donors result in better outcomes^1,22^ and that transplants involving MUDs and peripheral blood stem cell transplants are more likely to have greater magnitude of T cell proliferation (reviewed in by Dekker et al)^23^. The use of set correspondence theory principles to compare the entire telomere spectrum across donor and recipients, and alternatively the use of AUC measurement to arrive at a more robust measure of change across the entire span of TL measured, gives greater confidence in setting the stage for analysis.

Due to the limitations of this analysis, including its retrospective design and the relatively small patient cohort, these findings should be validated in a larger patient population. Ideally, this validation would involve measuring TL in both reconstituting myeloid and lymphoid populations, within a uniformly treated population. Uniformity in terms of donor selection, graft source, GVHD prophylaxis, and diagnosis for which transplantation was performed would also be desirable. It is also plausible that in patients undergoing GVHD prophylaxis using post-transplant cyclophosphamide, the TL attrition variability observed in these traditionally managed patients may be mitigated. Nevertheless, it will be of great clinical value if a lower delta TL in these patients correlates with relapse, providing an early biomarker to guide further cellular therapy, suc has DLI.

In conclusion, a detailed analysis of numerical data for telomere PCR amplification band lengths obtained by the TeSLA technique reveals that telomere length distribution across the human genome is mathematically ordered. The degree of TL attrition over the first 90 days following transplant is predictive of survival. This makes TL an important assay to predict transplant outcomes early in the course and devise corrective plans. Using simple mathematical principles to account for the non-linear nature of TL in the human genome yields clinically actionable information when compared to simpler methods which may not completely reflect the nature of complex biologic variables.

## Data Availability

All data produced in the present study are available upon reasonable request to the authors

## Acknowledgements.

The study is funded by the NIH grant U01AG066529 (The Telomere Research Network), and by the intramural program of the National Cancer Institute, NIH. The Center for International Blood and Marrow Transplant Research (CIBMTR) is supported primarily by Public Health Service U24CA076518 from the NCI, the National Heart, Lung and Blood Institute and the National Institute of Allergy and Infectious Diseases; HHSH250201700006C from the Health Resources and Services Administration; and N00014-21-1-2954 and N00014-20-1-2832 from the Office of Naval Research. Support for this study was provided by grants U10HL069294 and U24HL138660 to the Blood and Marrow Transplant Clinical Trials Network (BMT CTN) from the National Heart, Lung, and Blood Institute and the National Cancer Institute. T.P.L work was supported by NSF grant 2032119, NIH grants 1U01AG066529, 3U01AG066529-02S1, NCI contract 75N91019P00829, and New Jersey Alliance for Clinical and Translational Science Career Development Award NJACTS KL2 TR003018. The manuscript was prepared using BMT CTN 1202 Research Materials obtained from the BMT CTN Repository operated by the NMDP and does not necessarily reflect the opinions or views of the BMT CTN 1202 protocol team, the BMT CTN, the NHLBI, or NCI. We gratefully thank the institutions, investigators, and study participants. The authors also gratefully acknowledge Dr.s John Hansen, Peter Westervelt, Amin Alousi, Asad Bashey and Greg Yanik from the Fred Hutchinson Cancer Reseaerch Center, MD Anderson cancer Center, Northside Hospital, Barnes Jewish Hospita and University of Michigan for contributing patients to this study. The research use of blood samples and clinical Information was approved by the National Marrow Donor Program IRB. All study participants provided written informed consent for participation in the BMT-CTN 1202 protocol (NCT01879072) and the CIBMTR repository and database protocols (NCT00495300, and NCT01166009, respectively).

## Supplementary Tables and Figures

**Supplementary Table 1.** Characteristics of patient and HCT-related factors

| Variables | N (%) / Median (range) |
| --- | --- |
| Recipients' age at HCT (years) | 55.6 (3.3-73.9) |
| Donors' age | 34.3 (14.7-73.7) |
| Donors' sex |  |
| *Male* | 36 (50.0) |
| *Female* | 36 (50) |
| Recipients' sex |  |
| *Male* | 44 (61.1) |
| *Female* | 28 (38.9) |
| Disease |  |
| *Myeloid neoplasms* | 46 (63.9) |
| *Lymphoid neoplasms* | 19 (26.4) |
| *Other* | 7 (9.7) |
| Donors' type |  |
| *Matched sibling* | 25 (34.7) |
| *Haploidentical* | 5 (6.9) |
| *Unrelated donor* | 42 (58.3) |
| Stem cell source |  |
| *Bone marrow* | 14 (19.4) |
| *Peripheral blood* | 58 (80.6) |
| Conditioning regimen |  |
| *Myeloablative* | 45 (62.5) |
| *Nonmyeloablative* | 7 (9.7) |
| *Reduced intensity* | 20 (27.8) |
| *TBI-containing regimens* |  |
| *Yes* | 21 (29.2) |
| *No* | 51 (70.8) |

**Supplementary Table 2:** Distribution of delta TL by Geo delta, TL, Median delta-TL, and original delta-TL

|  | Mean (BP) | SD |
| --- | --- | --- |
| Geo delta-TL | 641 | 437 |
| Median delta-TL | 525 | 529 |
| Original delta-TL | 476 | 344 |

**Supplementary Table 3:**
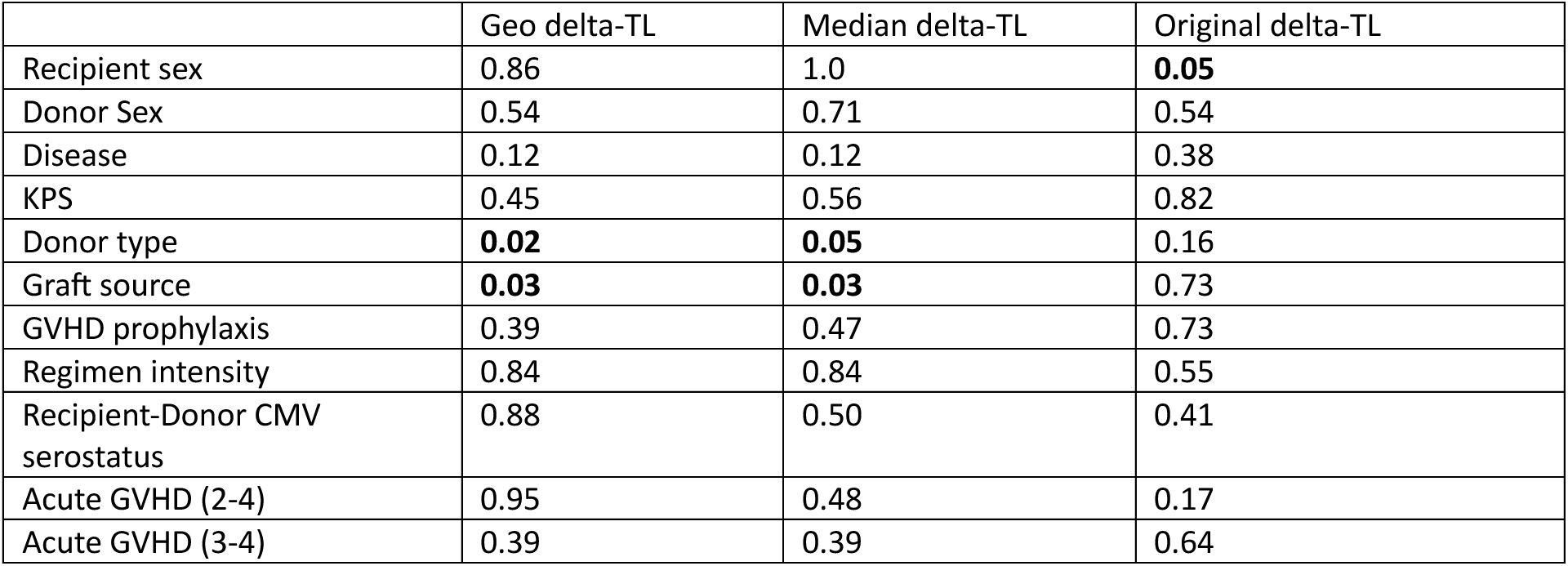
P-values for factors tested for associations with delta-TL by method of calculation

|  | Geo delta-TL | Median delta-TL | Original delta-TL |
| --- | --- | --- | --- |
| Recipient sex | 0.86 | 1.0 | **0.05** |
| Donor Sex | 0.54 | 0.71 | 0.54 |
| Disease | 0.12 | 0.12 | 0.38 |
| KPS | 0.45 | 0.56 | 0.82 |
| Donor type | **0.02** | **0.05** | 0.16 |
| Graft source | **0.03** | **0.03** | 0.73 |
| GVHD prophylaxis | 0.39 | 0.47 | 0.73 |
| Regimen intensity | 0.84 | 0.84 | 0.55 |
| Recipient-Donor CMV serostatus | 0.88 | 0.50 | 0.41 |
| Acute GVHD (2-4) | 0.95 | 0.48 | 0.17 |
| Acute GVHD (3-4) | 0.39 | 0.39 | 0.64 |

**Supplementary Table 4.** Adjusted hazard ratio of all-cause mortality in relation to post-HCT telomere length attrition calculated by different delta-TL methods

|  | Geo delta-TL |  | Median delta-TL |  | Original delta-TL |  |
| --- | --- | --- | --- | --- | --- | --- |
|  | HR (95% CI) | P |  |  | HR (95% CI) |  |
| Least shortening (Q1) | 2.53 (0.85-7.48) | 0.09 | 2.60 (0.91-7.48) | 0.07 | 0.99 (0.36-2.79) | 0.99 |
| Most shortening (Q4) | 1.72 (0.54-5.49) | 0.36 | 1.77 (0.55-5.66) | 0.34 | 0.94 (0.31-2.84) | 0.92 |
| Intermediate shortening (Q2, Q3) | Ref |  | Ref |  | Ref |  |

**Supplementary Table 5.** Frequency of causes of death by AUC delta-TL

|  | Least shortening (<25<sup>th</sup> percentile) | Most shortening (>75<sup>th</sup> percentile) | Intermediate (25-75<sup>th</sup> percentile) | Total |
| --- | --- | --- | --- | --- |
|  | N (%) |  |  |  |
| GvHD | 1 (20) | 3 (60) | 1 (20) | 5 |
| Organ Failure | 2 (50) | 1 (50) | 0 (0) | 3 |
| Relapse | 7 (58.3) | 3 (25) | 2 (16.7) | 12 |
| Unknown/other | 2 (100%) | 0 | 0 | 2 |
| Total | 12 | 7 | 3 | 22 |

**Supplementary figure 1.**
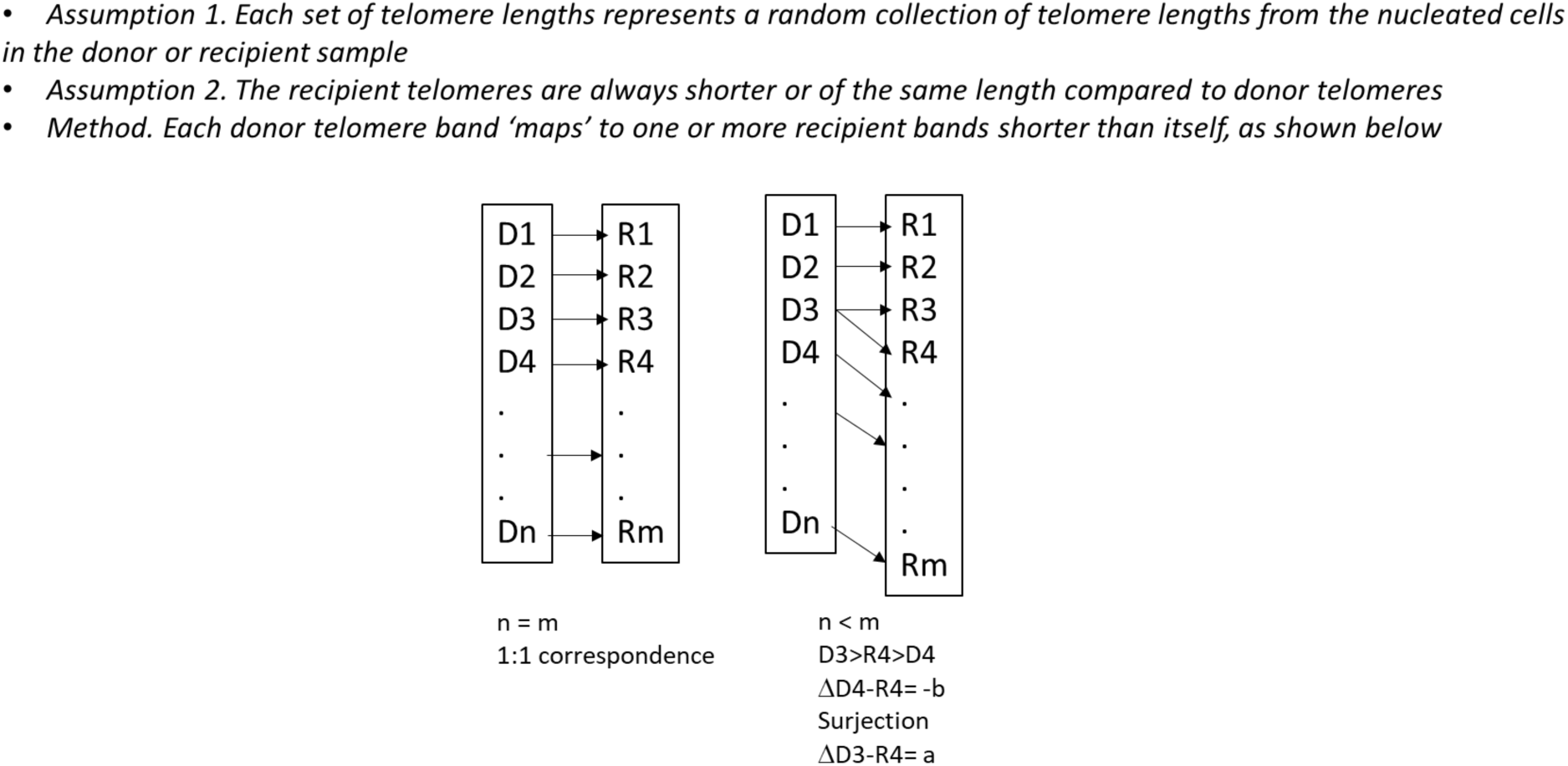
Set correspondence method illustrated.

**Supplementary Figure 2.**
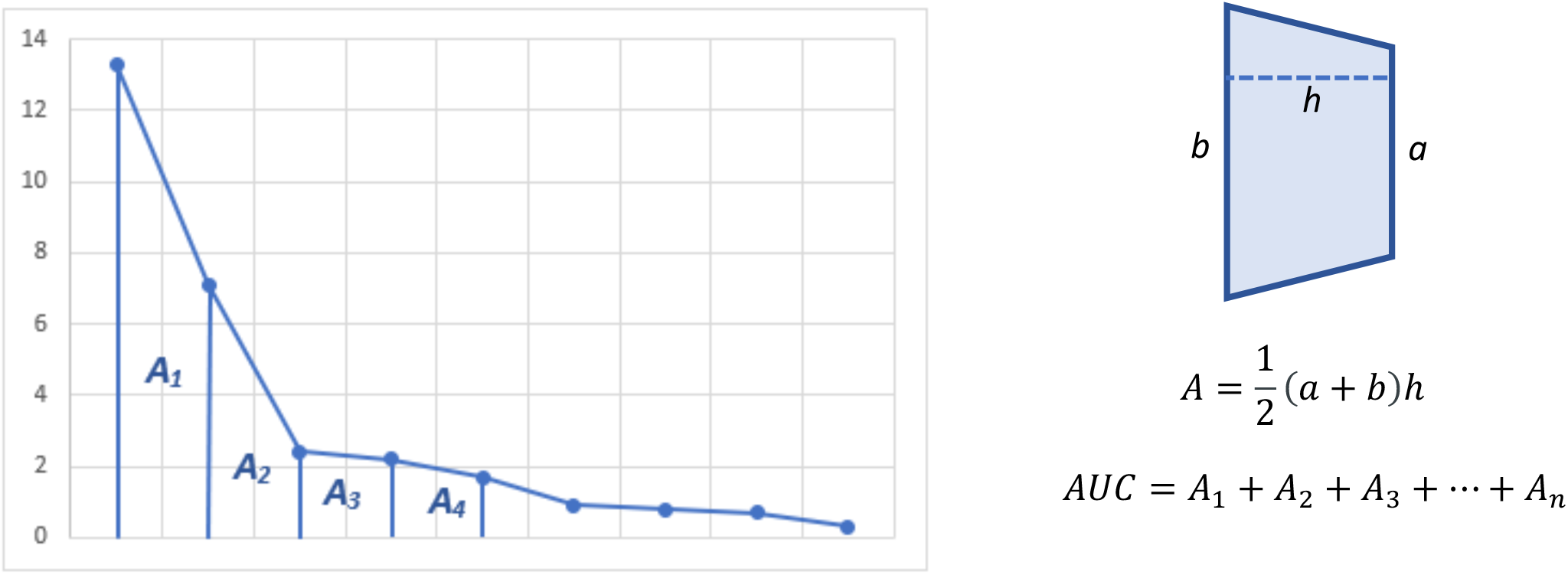
AUC calculation method illustrated.

**Supplementary Figure 3.**
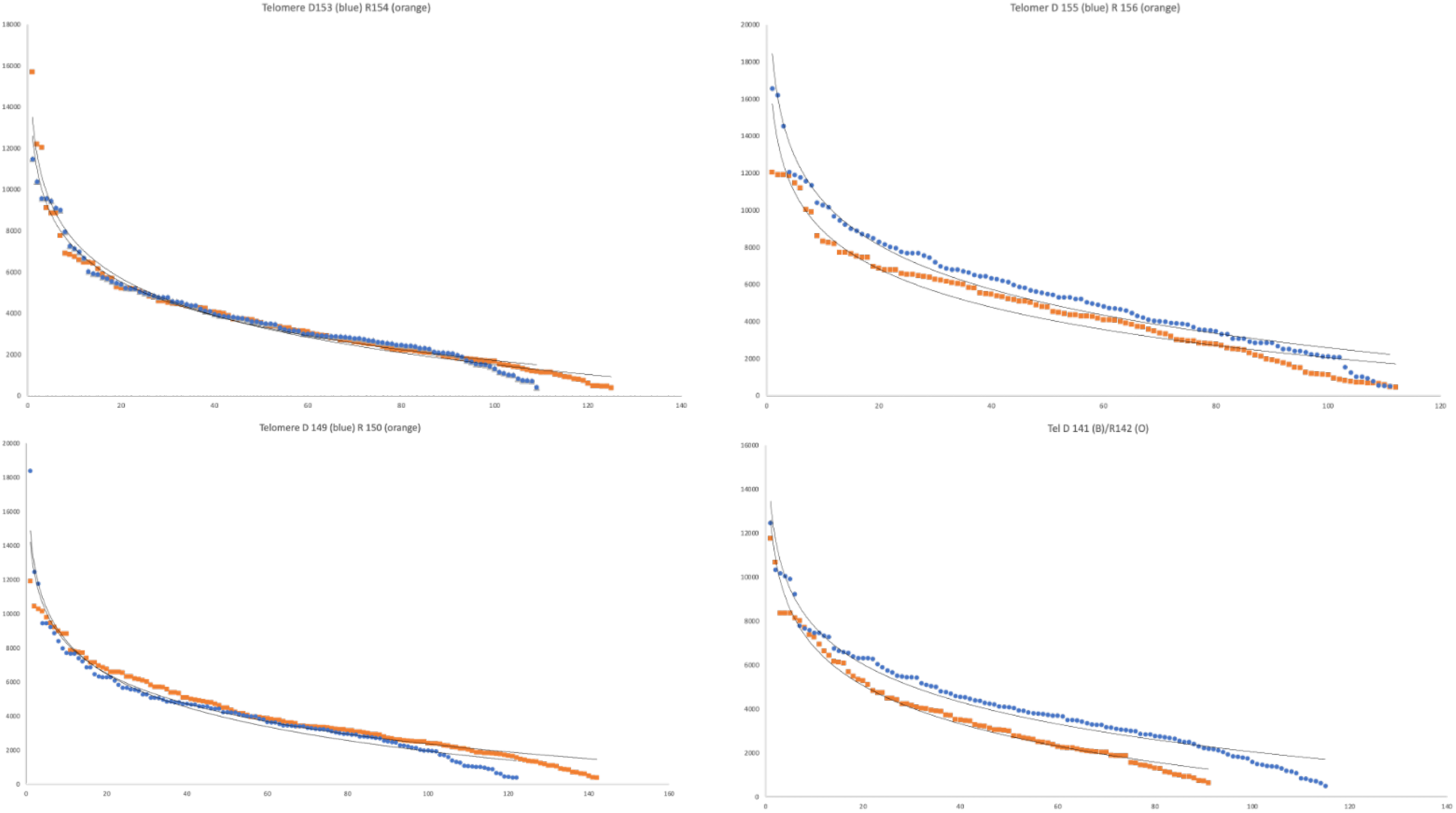
Variability in TL curves between transplant donors (blue) and recipients (orange). Successive band lengths (bp) depicted on the X axis.

**Supplementary Figure 4:**
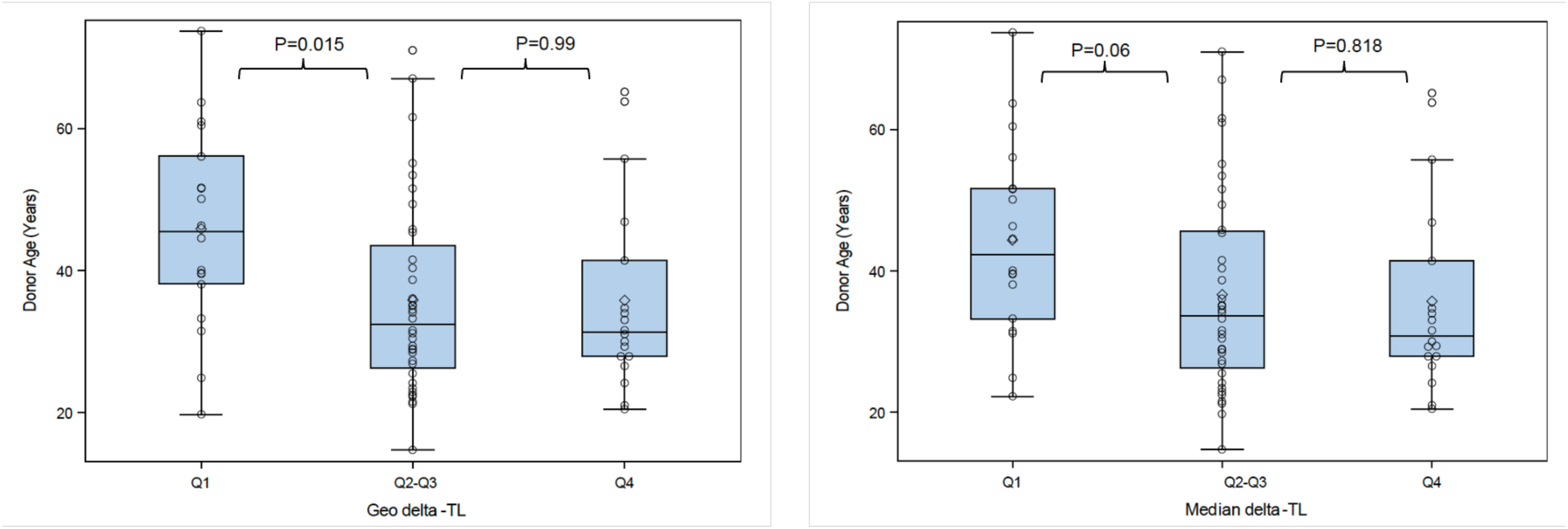
Relationship between donor age and delta-TL of different methods

## Appendix I

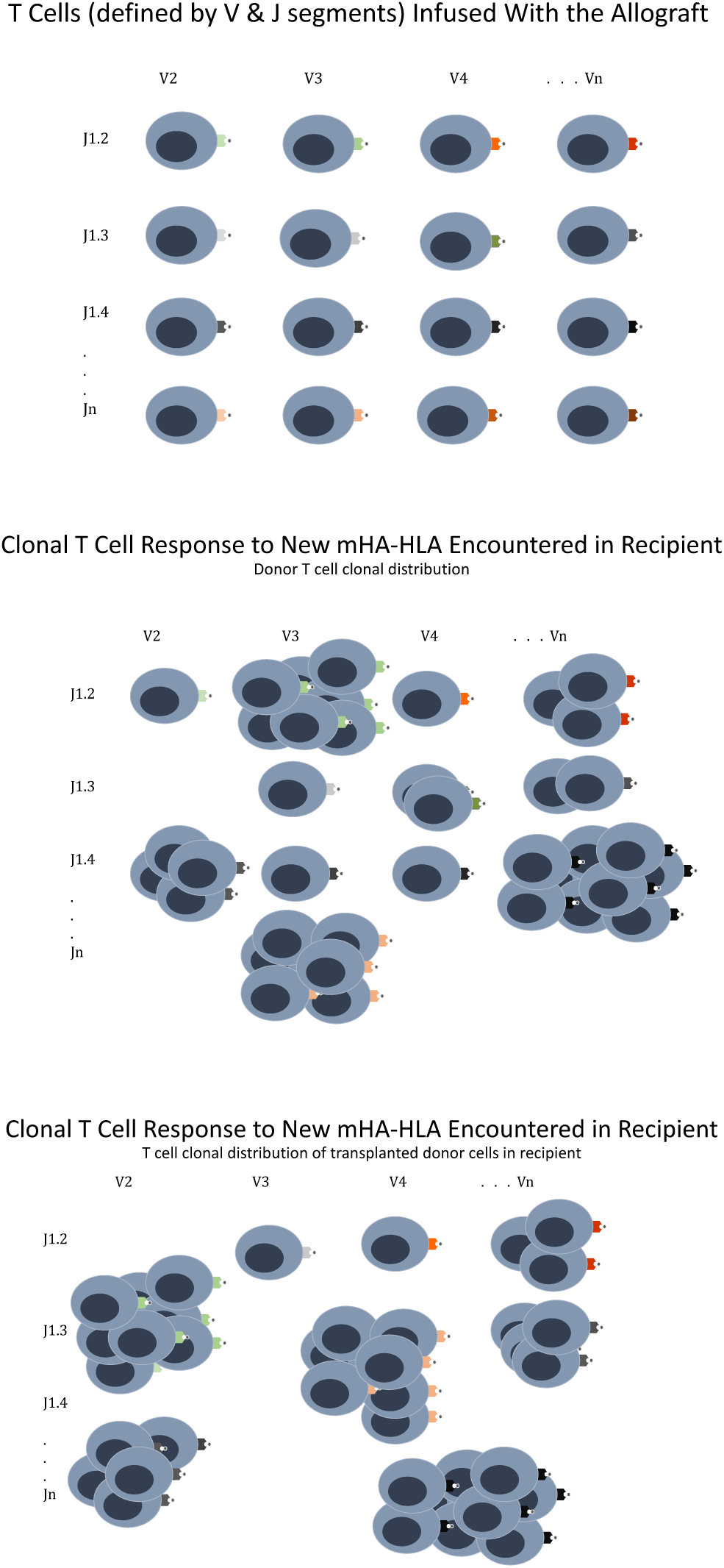

## Appendix II

Methodologically, ordered TL bands for donors and recipients were displayed after fitting growth equations,

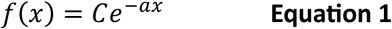

where *C* a constant and represents the Y-intercept of the growth curve, *a* the exponent represents the index by which the band length declines, *x* represents the (bands) or iterations of growth in this ascending order model, increases in value. This last variable *x* is measured as bands of different telomere length in the assay. The TL-AUC for both donors and recipients may be calculated as follow:

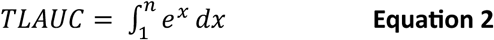

Difference between successive telomere band lengths plotted against the length of the entire telomere demonstrates an approximate log-periodic decline (inset). The difference in length between successive bands declines as a function of the length of the telomeres. When examined in descending order with an exponential decay model explored (**Figure X1**), the TL decay rate (exponent of the decay curve) was remarkably consistent across all the samples (**Figure X2**). In this paper the TL from a group of transplant donors and recipients is analyzed to demonstrate that the TL are not random, rather these follow an logarithmic distribution, and the difference in length between successive bands is determined by a consistent index.

**Figure X.**
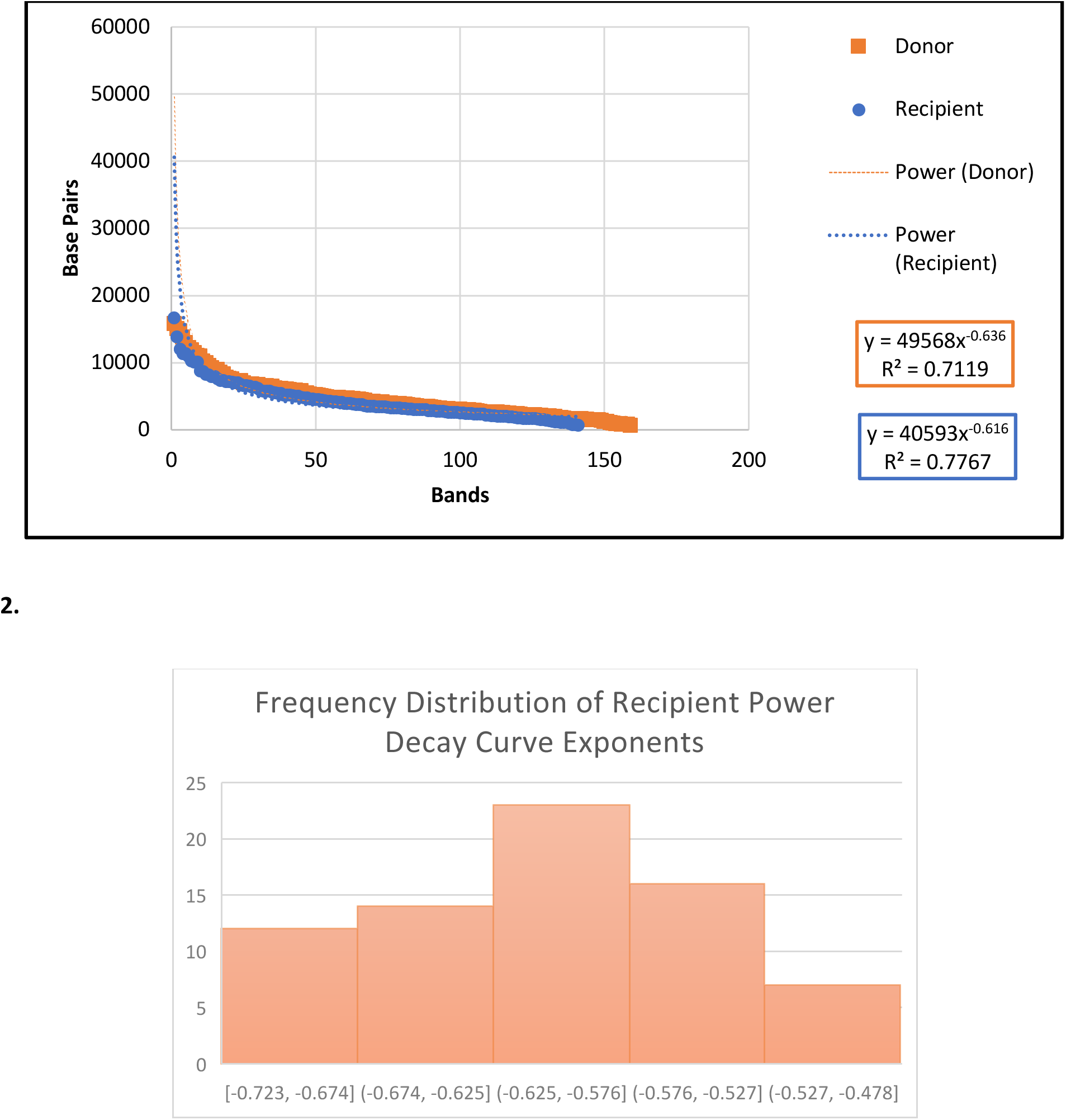
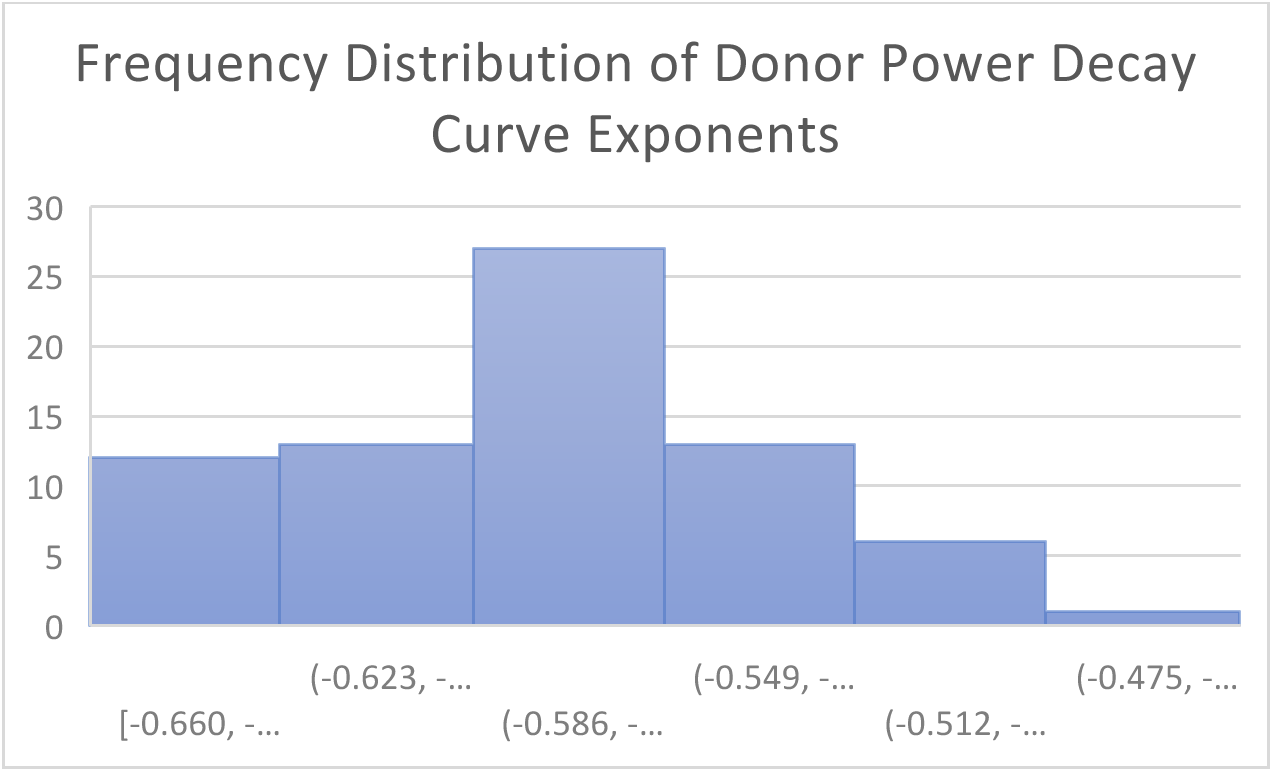
(**1**) Growth curve of telomere lengths with exponents calculated (Donor-blue; Recipient-orange). (**2**) Frequency of the exponents of the growth curve in different donors and recipients plotted.

### Set correspondence methodology

The surjection function employed to account for physiologic inconsistencies in the above model did not significantly alter the pattern of TL differences observed in each donor recipient pair (**Figure Y1 & Y2**), allowing the use of numerically simpler positive values of the surjected Di-Rj values. The median values of surjected geometric means of the Di-Rj for individual DRP was 574 base pairs (74 to 2055), while the median values of the Di-Rj medians for individual DRP was 559 (−710 to 1956), with frequency distribution consistent with disparate donor-recipient telomere length change over the first 100 days post-transplant (**Figure Z1 & Z2**).

**Figure Y.**
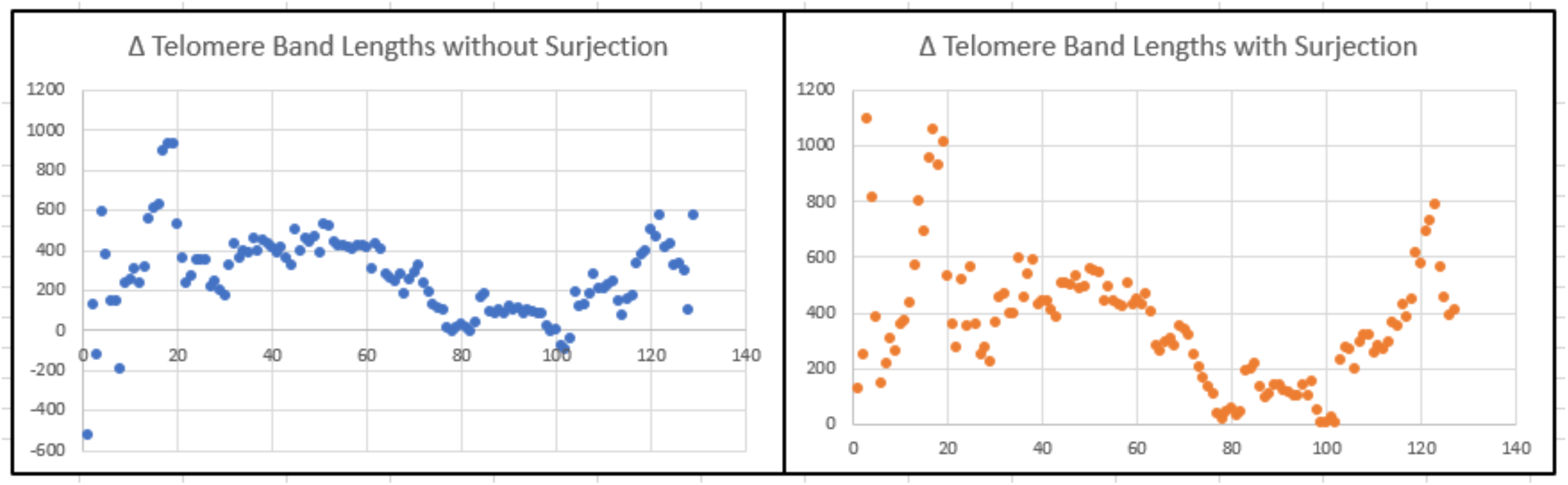
**(1)**. Representative graphs of a single DRP displaying the DDi-Rj with 1:1 correspondence and with surjection (surj). X-Axis has the successive *i^th^* and *j^th^* band, and Y axis depicts the difference between these in base pairs. As can be seen there are negative values in the graph to the left, indicating sporadic longer telomere band lengths in recipient.

**Figure Y.**
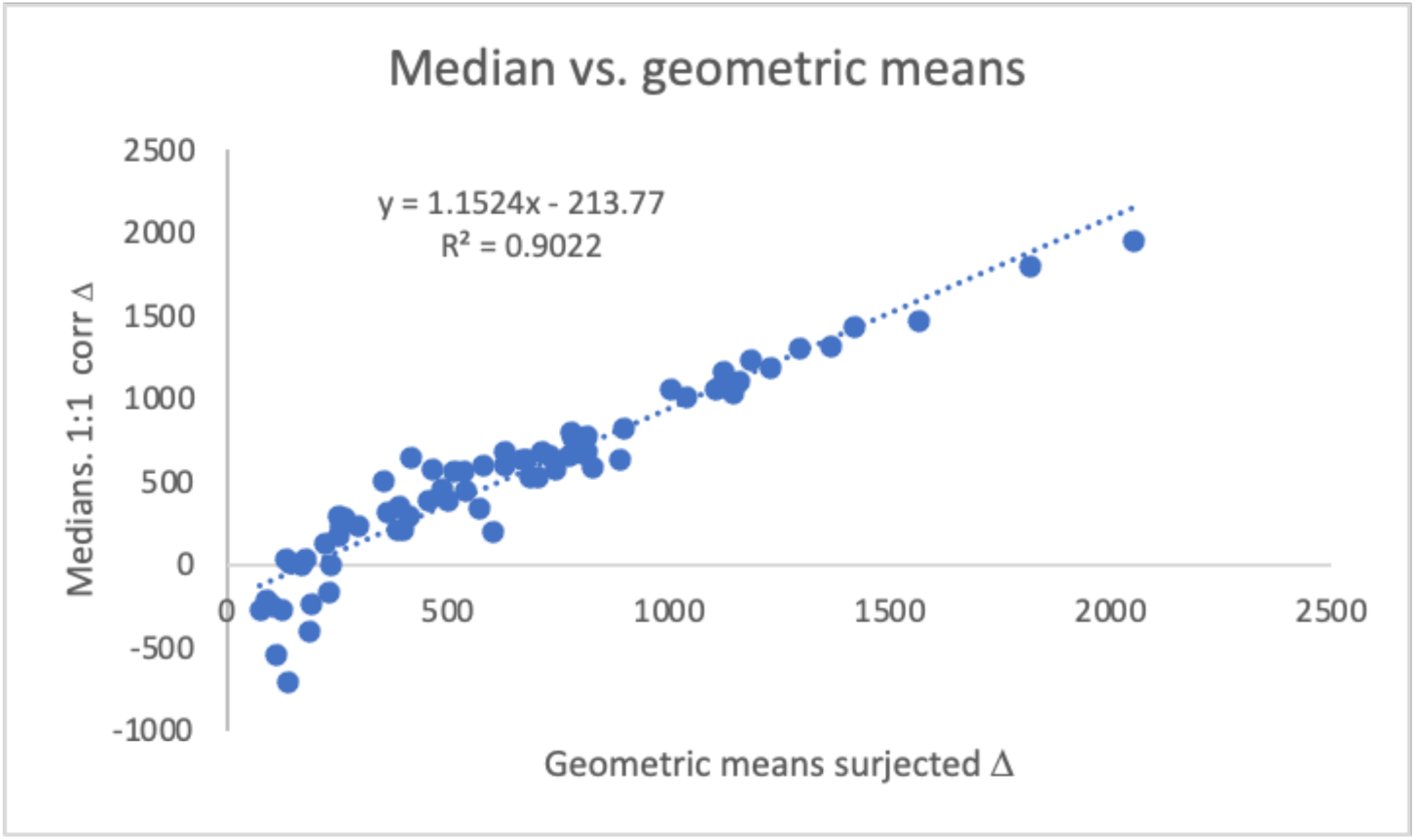
**(2)**. Correlation between surjected geometric mean and medians of the 1:1 correspondence sets for individual DRP (*P <0.00001*). Medians were used because the presence of 0 and negative values prevented geometric mean being calculated in the correspondence without surjection.

**Figure Z.**
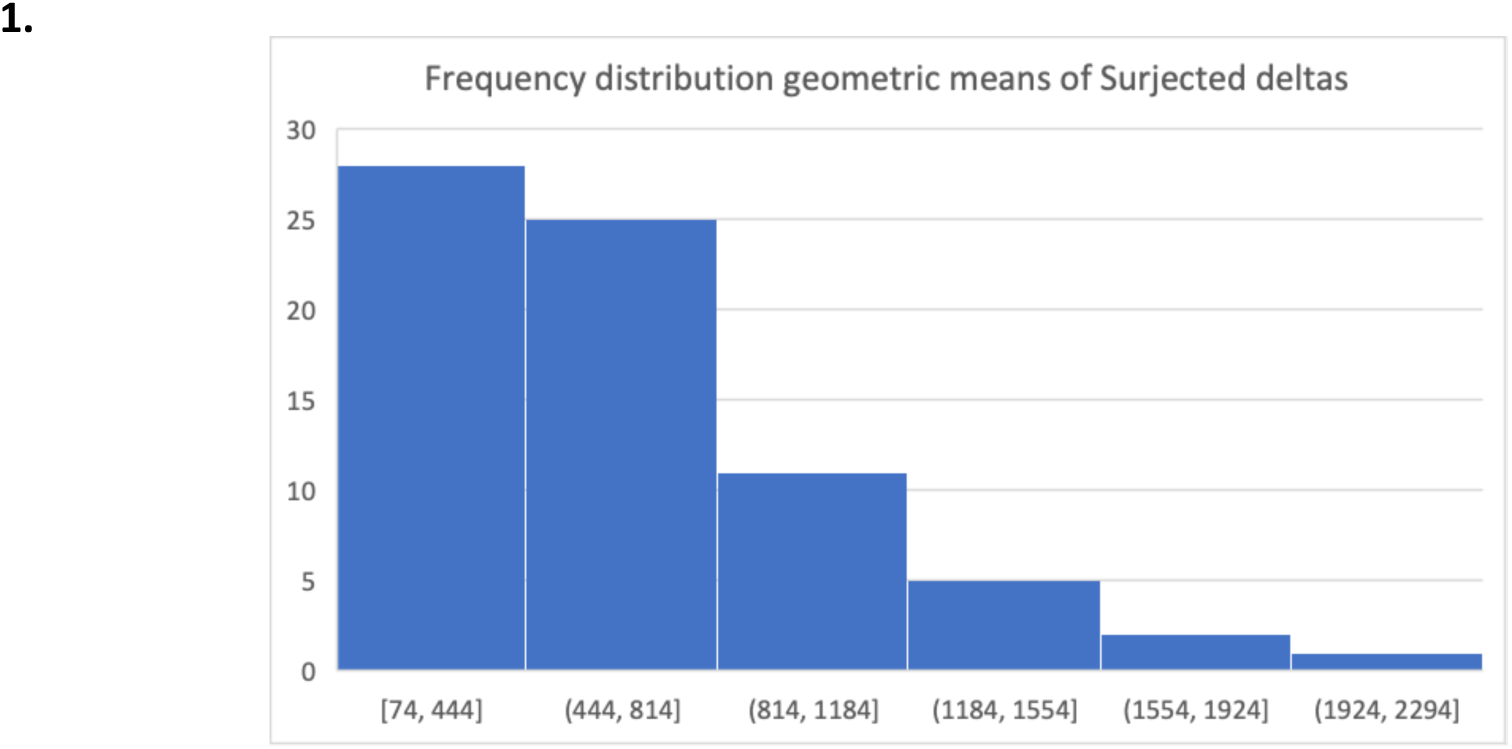
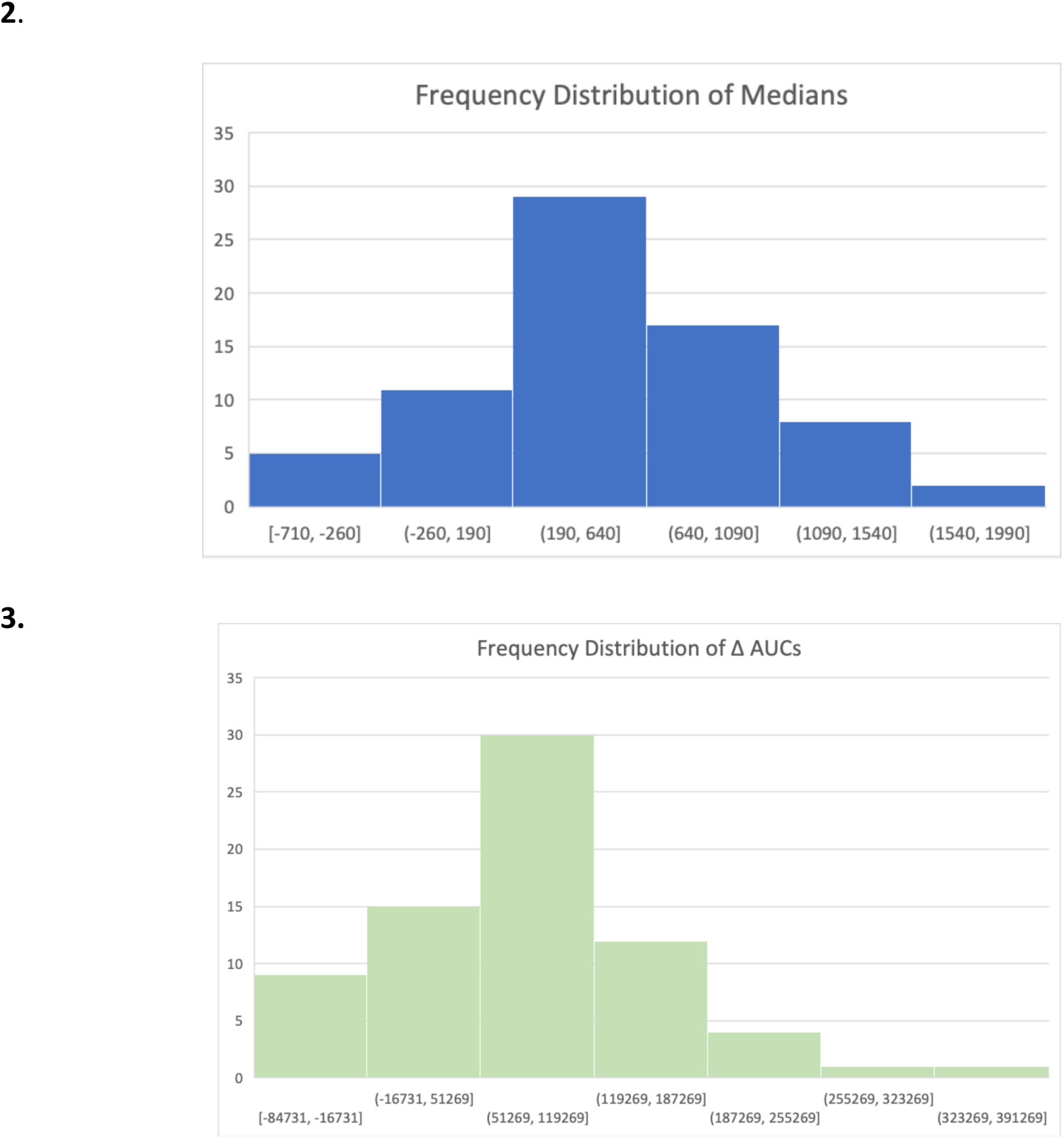
**(1).** Frequency distribution of surjected Di-Rj for each DRP, and (**2**) Di-Rj with 1:1 correspondence (3) Di-Rj with Δ AUC-TL.

**Figure W.**
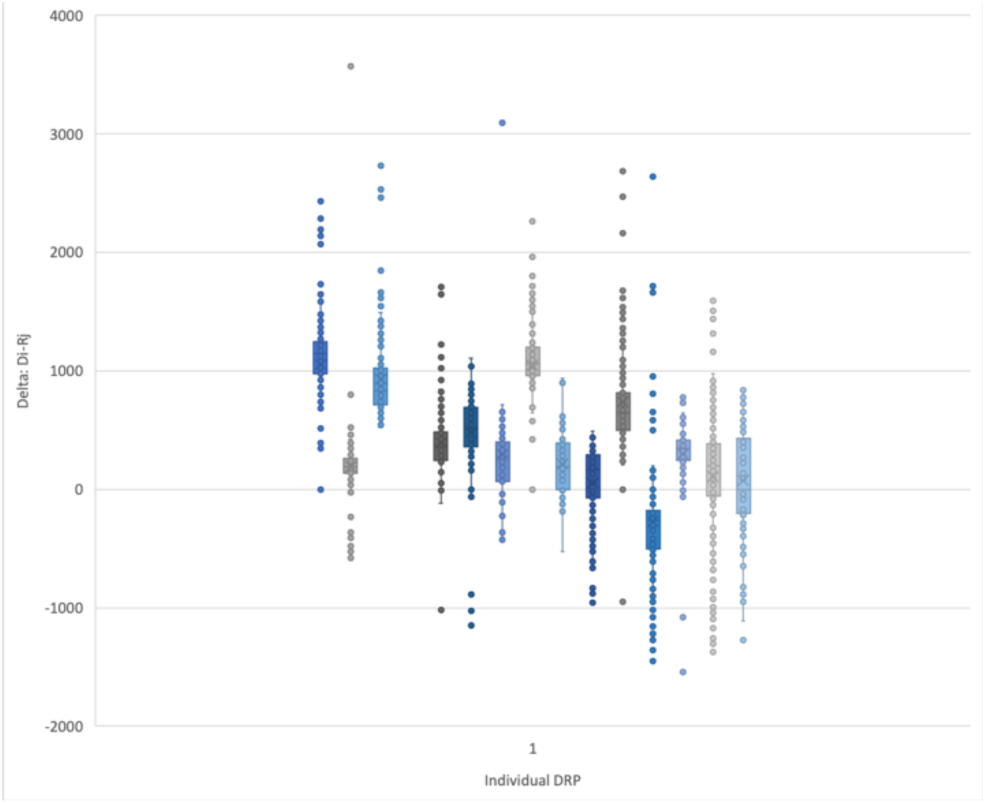
Plot demonstrating the variability of Di-Rj values for 15 donor-recipient pairs (DRP). Measured in base pairs.

## Footnotes

1 O’Connor, C. (2008) Telomeres of human chromosomes. *Nature Education* 1(1):166

3 https://www.mathworks.com/help/matlab/ref/trapz.html#bua4lsr

## References.

1. Kollman C, Spellman SR, Zhang M-J, et al. The effect of donor characteristics on survival after unrelated donor transplantation for hematologic malignancy. *Blood*, The Journal of the American Society of Hematology 2016; 127(2): 260–7.

2. Alousi AM, Le-Rademacher J, Saliba RM, et al. Who is the better donor for older hematopoietic transplant recipients: an older-aged sibling or a young, matched unrelated volunteer? *Blood*, The Journal of the American Society of Hematology 2013; 121(13): 2567–73.

3. Buhler S, Bettens F, Dantin C, et al. Genetic T-cell receptor diversity at 1 year following allogeneic hematopoietic stem cell transplantation. Leukemia 2020; 34(5): 1422–32.

4. Watkins B, Horan J, Storer B, Martin P, Carpenter P, Flowers M. Recipient and donor age impact the risk of developing chronic GvHD in children after allogeneic hematopoietic transplant. Bone marrow transplantation 2017; 52(4): 625–6.

5. Mehta J, Gordon L, Tallman M, et al. Does younger donor age affect the outcome of reduced-intensity allogeneic hematopoietic stem cell transplantation for hematologic malignancies beneficially? Bone marrow transplantation 2006; 38(2): 95–100.

6. Vescovini R, Fagnoni FF, Telera AR, et al. Naïve and memory CD8 T cell pool homeostasis in advanced aging: impact of age and of antigen-specific responses to cytomegalovirus. Age 2014; 36: 625–40.

7. Azuma E, Hirayama M, Yamamoto H, Komada Y. The role of donor age in naive T-cell recovery following allogeneic hematopoietic stem cell transplantation: the younger the better. Leukemia & lymphoma 2002; 43(4): 735–9.

8. Karimian K, Groot A, Huso V, et al. Human telomere length is chromosome end–specific and conserved across individuals. Science 2024; 384(6695): 533–9.

9. Baerlocher GM, Rovó A, Müller A, et al. Cellular senescence of white blood cells in very long-term survivors after allogeneic hematopoietic stem cell transplantation: the role of chronic graft-versus-host disease and female donor sex. *Blood*, The Journal of the American Society of Hematology 2009; 114(1): 219–22.

10. Akiyama M, Hoshi Y, Sakurai S, Yamada H, Yamada O, Mizoguchi H. Changes of telomere length in children after hematopoietic stem cell transplantation. Bone Marrow Transplantation 1998; 21(2): 167–71.

11. Akiyama M, Asai O, Kuraishi Y, et al. Shortening of telomeres in recipients of both autologous and allogeneic hematopoietic stem cell transplantation. Bone Marrow Transplantation 2000; 25(4): 441–7.

12. Lai T-P, Wright WE, Shay JW. Comparison of telomere length measurement methods. Philosophical Transactions of the Royal Society B: Biological Sciences 2018; 373(1741): 20160451.

13. Toor AA, Sabo RT, Roberts CH, et al. Dynamical system modeling of immune reconstitution after allogeneic stem cell transplantation identifies patients at risk for adverse outcomes. Biology of Blood and Marrow Transplantation 2015; 21(7): 1237–45.

14. Koparde V, Abdul Razzaq B, Suntum T, et al. Dynamical system modeling to simulate donor T cell response to whole exome sequencing-derived recipient peptides: Understanding randomness in alloreactivity incidence following stem cell transplantation. PLoS One 2017; 12(12): e0187771.

15. Razzaq BA, Scalora A, Koparde VN, et al. Dynamical System Modeling to Simulate Donor T Cell Response to Whole Exome Sequencing-Derived Recipient Peptides Demonstrates Different Alloreactivity Potential in HLA-Matched and-Mismatched Donor–Recipient Pairs. Biology of Blood and Marrow Transplantation 2016; 22(5): 850–61.

16. Lai T-P, Zhang N, Noh J, et al. A method for measuring the distribution of the shortest telomeres in cells and tissues. Nature Communications 2017; 8(1): 1356.

17. Baird DM, Rowson J, Wynford-Thomas D, Kipling D. Extensive allelic variation and ultrashort telomeres in senescent human cells. Nature Genetics 2003; 33(2): 203–7.

18. Lai TP, Verhulst S, Dagnall CL, Hutchinson A, Spellman SR, Howard A, Katki HA, Levine JE, Saber W, Aviv A, Gadalla SM. Decoupling blood telomere length from age in recipients of allogeneic hematopoietic cell transplant in the BMT-CTN 1202. Front Immunol. 2022 Oct 3;13:966301. doi: 10.3389/fimmu.2022.966301. PMID: 36263045; PMCID: PMC9574912.

19. Bolon YT, Atshan R, Allbee-Johnson M, Estrada-Merly N, Auletta JJ, Broglie L, Cusatis R, Page KM, Phelan R, Sajulga R Jr, Shaw BE, Spahn A, Steinert P, Stewart V, Vierra-Green C, Lee SJ, Spellman SR. Leveraging Hematopoietic Cell Transplant Data and Biorepository Resources at the Center for International Blood and Marrow Transplant Research to Improve Patient Outcomes. Transplant Cell Ther. 2024 Sep;30(9):921.e1-921.e22. doi: 10.1016/j.jtct.2024.06.010. Epub 2024 Jun 11. PMID: 38871054.

20. Reshef R, Saber W, Bolaños-Meade J, Chen G, Chen YB, Ho VT, Ponce DM, Nakamura R, Martens MJ, Hansen JA, Levine JE. Acute GVHD Diagnosis and Adjudication in a Multicenter Trial: A Report From the BMT CTN 1202 Biorepository Study. J Clin Oncol. 2021 Jun 10;39(17):1878–1887. doi: 10.1200/JCO.20.00619. Epub 2021 Jan 28. PMID: 33507810; PMCID: PMC8260916.

21. Helby J, Petersen SL, Kornblit B, Nordestgaard BG, Mortensen BK, Bojesen SE, Sengeløv H. Mononuclear cell telomere attrition is associated with overall survival after nonmyeloablative allogeneic hematopoietic cell transplantation for hematologic malignancies. Biology of Blood and Marrow Transplantation 2019; 25(3): 496–504.

22. Shaw BE, Logan BR, Spellman SR, Marsh SGE, Robinson J, Pidala J, Hurley C, Barker J, Maiers M, Dehn J, Wang H, Haagenson M, Porter D, Petersdorf EW, Woolfrey A, Horowitz MM, Verneris M, Hsu KC, Fleischhauer K, Lee SJ. Development of an Unrelated Donor Selection Score Predictive of Survival after HCT: Donor Age Matters Most. Biol Blood Marrow Transplant. 2018 May;24(5):1049–1056. doi: 10.1016/j.bbmt.2018.02.006. Epub 2018 Feb 14. PMID: 29454040; PMCID: PMC5953795.

23. Dekker L, de Koning C, Lindemans C, Nierkens S. Reconstitution of T Cell Subsets Following Allogeneic Hematopoietic Cell Transplantation. Cancers (Basel). 2020 Jul 20;12(7):1974. doi: 10.3390/cancers12071974. PMID: 32698396; PMCID: PMC7409323.

